# A Predictive Model for Severe Covid-19 in the Medicare Population: A Tool for Prioritizing Scarce Vaccine Supply

**DOI:** 10.1101/2020.10.28.20219816

**Authors:** Bettina Experton, Hassan A. Tetteh, Nicole Lurie, Peter Walker, Colin J. Carroll, Adrien Elena, Christopher S. Hein, Blake Schwendiman, Justin L. Vincent, Christopher R. Burrow

**Affiliations:** Project Salus healthcare analytics group at Humetrix Inc., Del Mar, CA; Department of Defense Joint Artificial Intelligence Center (JAIC) Warfighter Health Mission Team, Washington, D.C; Coalition for Epidemic Preparedness Innovation (CEPI), Oslo, and Harvard Medical School, Boston, MA; Johns Hopkins University Applied Physics Laboratory, Baltimore, MD; Amazon Web Services Inc., Seattle, WA

## Abstract

**Background:** Recommendations for prioritizing populations for COVID-19 vaccination have focused on front-line health care personnel and residents in long term care, followed by other individuals at higher risk for severe disease. Existing models for identifying higher risk individuals including those over age 65 lack the needed integration of socio-demographic and clinical risk factors to ensure equitable vaccine allocation.

**Methods:** We developed a predictive model for severe COVID-19 using clinical data from de-identified Medicare claims for 16 million Medicare fee-for-service beneficiaries, including 1 million COVID-19 cases, and socio-economic data from the CDC Social Vulnerability Index. To identify risk factors for severe COVID-19, we used multivariate logistic regression and random forest modeling. Predicted individual probabilities of COVID-19 hospitalization were then calculated for population risk stratification and COVID-19 vaccine prioritization, and for mapping of population risk levels at the county and zip code levels on a nationwide dashboard.

**Results:** The leading Covid-19 hospitalization risk factors driving the risk model were: Non-white ethnicity (particularly North American Native, Black, and Hispanic), end-stage renal disease, advanced age (particularly age over 85), prior hospitalization, leukemia, morbid obesity, chronic kidney disease, lung cancer, chronic liver disease, pulmonary fibrosis or pulmonary hypertension, and chemotherapy. However, previously reported risk factors such as chronic obstructive pulmonary disease and diabetes conferred modest hospitalization risk. Among all social vulnerability factors analyzed, residence in a low-income zip code was the only risk factor independently predicting Covid-19 hospitalization. The mapped hospitalization risk levels showed significant correlations with the cumulative COVID-19 case hospitalization rates at the zip code level in the fifteen most populous U.S. major metropolitan areas.

**Conclusion:** This multi-factor risk model which predicts severe Covid-19and its population risk dashboard can be used to optimize Covid-19 vaccine allocation in the higher risk Medicare population where socio-demographic and comorbidity risk factors need to be considered concurrently.

## INTRODUCTION

The question of who should get Covid-19 vaccines first has been debated by the National Academy of Medicine, the Centers for Disease Control and Prevention, and by epidemiologists and other disease experts worldwide. U.S. recommendations currently focus on vaccinating individuals at risk for severe disease once healthcare personnel, nursing home residents, and essential workers are vaccinated.^1^ Individuals over age 65, who are also Medicare beneficiaries, account for a disproportionate share of hospitalizations and 80% of total Covid-19 related deaths.^2^

The National Academy of Medicine recommends simultaneously prioritizing individuals living in areas with socioeconomic conditions associated with disproportionate vulnerability^3,4^ by using the Centers for Disease Control and Prevention (CDC) Social Vulnerability Index (SVI)^5^ or the Covid-19 Community Vulnerability Index (CCVI).^6^ Complicating any allocation scheme is the fact that risk categories overlap;^7^ more than half of the 53 million Americans over age 65 suffer from two or more chronic conditions,^8^ and many of them also live in socially vulnerable areas.

Any prioritization scheme, following on the Advisory Committee on Immunization Practices,^9^ must consider clinical, demographic and social vulnerability risks together. The current CDC listing of risk factors for severe Covid-19^10^ is derived from single hospital-based studies with limited sample sizes,^11,12^ or hospital reporting, both of which lack nationwide representation.^13^ The Center for Medicare and Medicaid Services (CMS) monthly Medicare Covid-19 Data Snapshots include national demographic characteristics and prevalence of common chronic conditions among hospitalized fee-for-service (FFS) Medicare beneficiaries, but these lack more detailed clinical and socio-economic data needed to identify or stratify at risk populations.^14^ We are not aware of any published analyses that fully support simultaneous prioritization using both clinical and social vulnerability data.

We developed a model to predict Covid-19 hospitalization and death for Medicare beneficiaries using de-identified Medicare claims and the data comprising the CDC SVI. While the initial impetus for the work, developed for the Department of Defense (DoD) Joint Artificial Intelligence Center (JAIC), was to provide logistics support to hospitals overwhelmed by the pandemic, this model can also support operationalization of the National Academy of Medicine (NAM) and CDC recommendations for a COVID-19 vaccination campaign by stratifying the population by risk, and by mapping locations of beneficiaries in different risk strata.^15^

## METHODS

### Data sources

We constructed an observational cohort consisting of all Medicare FFS beneficiaries who since January 1, 2020, either had a Covid-19 test or diagnosis, or for any medical reason were hospitalized or had an emergency department, urgent care, or telehealth visit. For this cohort, we received weekly data outputs from the Center for Medicare and Medicaid Services (CMS) Chronic Condition Warehouse from October 1^st^, 2019 through November 22^d^ 2020, resulting in a weekly download of over 100 million individual records to a secure government enclave of the DoD JAIC for Project Salus’ partner, Humetrix, to process and analyze.

### Dependent and Independent variables

Dependent variables were confirmed Covid-19 cases and their related hospitalizations and deaths. Independent variables included: beneficiary age, sex, ethnicity, insurance coverage, residential zip code. We used the number of prior hospital admissions since October 1, 2019 as an indicator of frailty, and we assessed comorbidities by examining individual’s diagnoses listed in claims data beginning October 1, 2019. To identify comorbidities, we used a set of chronic conditions flagged by CMS and compiled diagnostic categories using specific ICD-10 code algorithms to identify additional chronic conditions. Medication NDC codes were used to identify active pharmaceutical ingredients, which were grouped by pharmaceutical class by mapping to RxNorm codes. Socio-economic variables (e.g., income, housing, and other factors) were defined at the individual residential zip code level after conversion from the census track based data found in the CDC Social Vulnerability Index using HUD-USPS^16^ crosswalk files. A complete list of independent variables is found in the Supplemental Appendix.

### Statistical analysis, variable selection and risk model

We used logistic regression to identify significant predictors of Covid-19 related hospitalization or Covid-19 deaths, (R statistical software, version 3.6 with rms, glmnet and pROC packages),^17-20^ using the following binary outcomes: those who received outpatient care only (defined as cases that did not require hospitalization or didn’t die at least thirty days after diagnosis) versus either those who were hospitalized for Covid-19, or those beneficiaries whose deaths were attributed to Covid-19 (defined as cases who died of SARS-CoV-2 infection within 60 days of diagnosis). We divided our sample into training (60%) and validation (40%) sets to develop our final models, randomly allocating cases to training or validation components in a 50:50 ratio. We examined correlation coefficients between independent variables and used lasso regression to eliminate correlated or collinear independent variables. We then used stepwise backward variable selection procedure based on the Akaike Information Criterion (AIC) to remove non-significant variables.

Because the odds ratios derived from logistic regression models are a measure of the association between a given feature (e.g. North American Native ethnicity) and the outcome (e.g. hospitalization), we supplemented our analyses with a random forest machine learning algorithm, which produces computed Feature Importance values (Python, scikit-learn version 0.22.1 with RandomForestClassifier and GridSearchCV packages)^21^ and provides information about the relative importance of each feature for predicting outcomes for the entire sample. We calculated the Gini importance of each feature in the models to determine which variables were the most important for determining severe disease outcomes in our sample. The data sampling procedure, variable definition, feature engineering, and patient outcome definitions were identical to those described above for Logistic Regression.

Statistical analysis details are provided in the online Supplement.

### Population level Covid-19 hospitalization risk mapping

We used the results of our logistic regression to compute individual predicted probabilities of hospitalization in the event of SARS-CoV-2 infection for the entire analytic cohort of 16 million beneficiaries. We then computed the percentage of the cohort population with a predicted probability of hospitalization of 0.55 or higher, mapping the data at county and zip code levels for the entire country on a nationwide digital dashboard.

## RESULTS

### Study population characteristics

Socio-demographic and clinical characteristics of the study population (16 million beneficiaries with 1,030,893 confirmed Covid-19 cases, as of November 20, 2020) are summarized in Table 1.

**Table 1.**
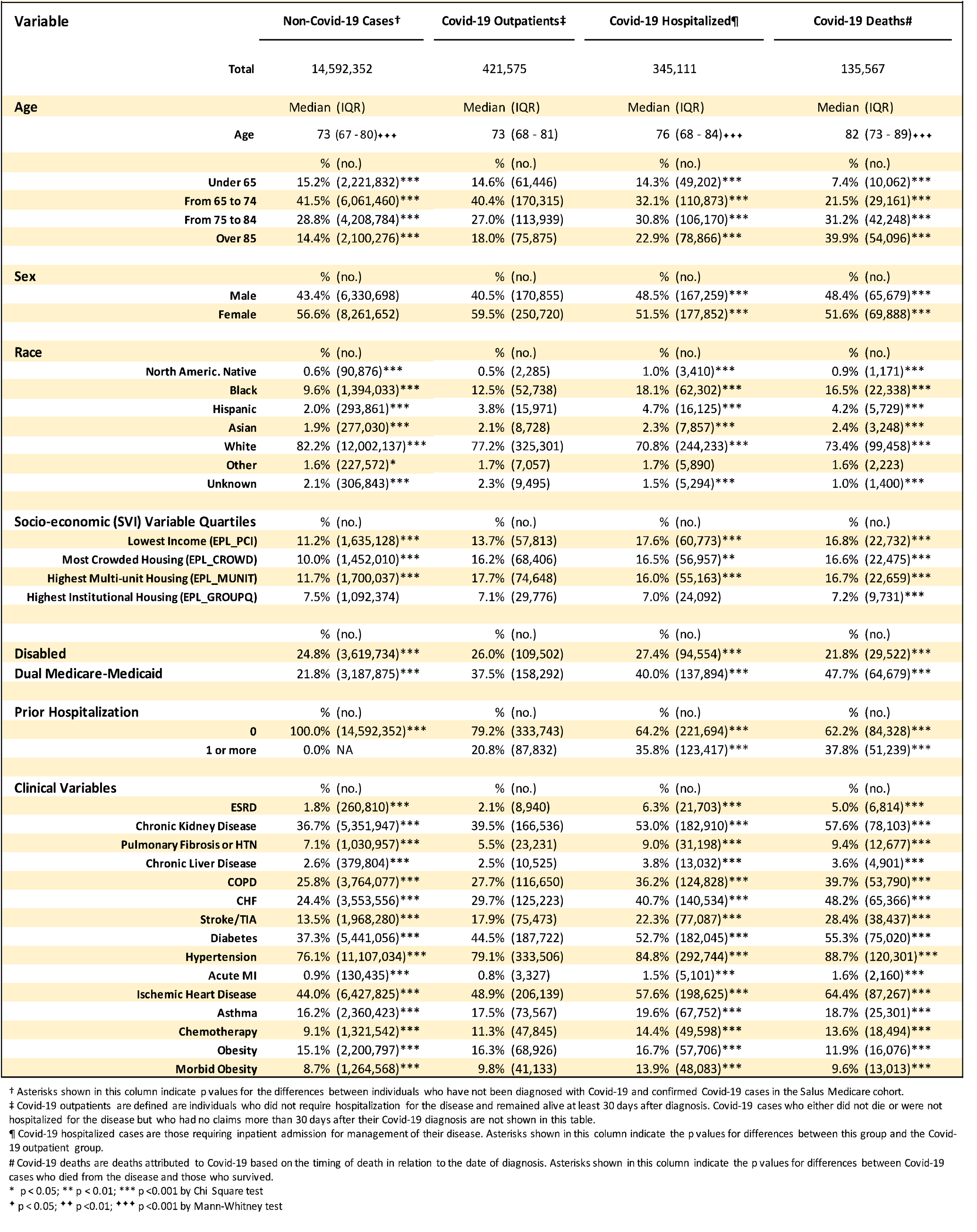
Demographic, Clinical and Socioeconomic Characteristics of the Project Salus Medicare Cohort.

| Variable | Non-Covid-19 Cases† | Covid-19 Outpatients‡ | Covid-19 Hospitalized¶ | Covid-19 Deaths# |
| --- | --- | --- | --- | --- |
| **Total** | 14,592,352 | 421,575 | 345,111 | 135,567 |
| **Age** | Median (IQR) | Median (IQR) | Median (IQR) | Median (IQR) |
| Age | 73 (67 - 80)+++ | 73 (68 - 81) | 76 (68 - 84)+++ | 82 (73 - 89)+++ |
|  | % (no.) | % (no.) | % (no.) | % (no.) |
| Under 65 | 15.2% (2,221,832)*** | 14.6% (61,446) | 14.3% (49,202)*** | 7.4% (10,062)*** |
| From 65 to 74 | 41.5% (6,061,460)*** | 40.4% (170,315) | 32.1% (110,873)*** | 21.5% (29,161)*** |
| From 75 to 84 | 28.8% (4,208,784)*** | 27.0% (113,939) | 30.8% (106,170)*** | 31.2% (42,248)*** |
| Over 85 | 14.4% (2,100,276)*** | 18.0% (75,875) | 22.9% (78,866)*** | 39.9% (54,096)*** |
| **Sex** | % (no.) | % (no.) | % (no.) | % (no.) |
| Male | 43.4% (6,330,698) | 40.5% (170,855) | 48.5% (167,259)*** | 48.4% (65,679)*** |
| Female | 56.6% (8,261,652) | 59.5% (250,720) | 51.5% (177,852)*** | 51.6% (69,888)*** |
| **Race** | % (no.) | % (no.) | % (no.) | % (no.) |
| North Americ. Native | 0.6% (90,876)*** | 0.5% (2,285) | 1.0% (3,410)*** | 0.9% (1,171)*** |
| Black | 9.6% (1,394,033)*** | 12.5% (52,738) | 18.1% (62,302)*** | 16.5% (22,338)*** |
| Hispanic | 2.0% (293,861)*** | 3.8% (15,971) | 4.7% (16,125)*** | 4.2% (5,729)*** |
| Asian | 1.9% (277,030)*** | 2.1% (8,728) | 2.3% (7,857)*** | 2.4% (3,248)*** |
| White | 82.2% (12,002,137)*** | 77.2% (325,301) | 70.8% (244,233)*** | 73.4% (99,458)*** |
| Other | 1.6% (227,572)* | 1.7% (7,057) | 1.7% (5,890) | 1.6% (2,223) |
| Unknown | 2.1% (306,843)*** | 2.3% (9,495) | 1.5% (5,294)*** | 1.0% (1,400)*** |
| **Socio-economic (SVI) Variable Quartiles** | % (no.) | % (no.) | % (no.) | % (no.) |
| Lowest Income (EPL_PCI) | 11.2% (1,635,128)*** | 13.7% (57,813) | 17.6% (60,773)*** | 16.8% (22,732)*** |
| Most Crowded Housing (EPL_CROWD) | 10.0% (1,452,010)*** | 16.2% (68,406) | 16.5% (56,957)** | 16.6% (22,475)*** |
| Highest Multi-unit Housing (EPL_MUNIT) | 11.7% (1,700,037)*** | 17.7% (74,648) | 16.0% (55,163)*** | 16.7% (22,659)*** |
| Highest Institutional Housing (EPL_GROUPQ) | 7.5% (1,092,374) | 7.1% (29,776) | 7.0% (24,092) | 7.2% (9,731)*** |
| **Disabled** | % (no.) | % (no.) | % (no.) | % (no.) |
| Disabled | 24.8% (3,619,734)*** | 26.0% (109,502) | 27.4% (94,554)*** | 21.8% (29,522)*** |
| **Dual Medicare-Medicaid** | 21.8% (3,187,875)*** | 37.5% (158,292) | 40.0% (137,894)*** | 47.7% (64,679)*** |
| **Prior Hospitalization** | % (no.) | % (no.) | % (no.) | % (no.) |
| 0 | 100.0% (14,592,352)*** | 79.2% (333,743) | 64.2% (221,694)*** | 62.2% (84,328)*** |
| 1 or more | 0.0% NA | 20.8% (87,832) | 35.8% (123,417)*** | 37.8% (51,239)*** |
| **Clinical Variables** | % (no.) | % (no.) | % (no.) | % (no.) |
| ESRD | 1.8% (260,810)*** | 2.1% (8,940) | 6.3% (21,703)*** | 5.0% (6,814)*** |
| Chronic Kidney Disease | 36.7% (5,351,947)*** | 39.5% (166,536) | 53.0% (182,910)*** | 57.6% (78,103)*** |
| Pulmonary Fibrosis or HTN | 7.1% (1,030,957)*** | 5.5% (23,231) | 9.0% (31,198)*** | 9.4% (12,677)*** |
| Chronic Liver Disease | 2.6% (379,804)*** | 2.5% (10,525) | 3.8% (13,032)*** | 3.6% (4,901)*** |
| COPD | 25.8% (3,764,077)*** | 27.7% (116,650) | 36.2% (124,828)*** | 39.7% (53,790)*** |
| CHF | 24.4% (3,553,556)*** | 29.7% (125,223) | 40.7% (140,534)*** | 48.2% (65,366)*** |
| Stroke/TIA | 13.5% (1,968,280)*** | 17.9% (75,473) | 22.3% (77,087)*** | 28.4% (38,437)*** |
| Diabetes | 37.3% (5,441,056)*** | 44.5% (187,722) | 52.7% (182,045)*** | 55.3% (75,020)*** |
| Hypertension | 76.1% (11,107,034)*** | 79.1% (333,506) | 84.8% (292,744)*** | 88.7% (120,301)*** |
| Acute MI | 0.9% (130,435)*** | 0.8% (3,327) | 1.5% (5,101)*** | 1.6% (2,160)*** |
| Ischemic Heart Disease | 44.0% (6,427,825)*** | 48.9% (206,139) | 57.6% (198,625)*** | 64.4% (87,267)*** |
| Asthma | 16.2% (2,360,423)*** | 17.5% (73,567) | 19.6% (67,752)*** | 18.7% (25,301)*** |
| Chemotherapy | 9.1% (1,321,542)*** | 11.3% (47,845) | 14.4% (49,598)*** | 13.6% (18,494)*** |
| Obesity | 15.1% (2,200,797)*** | 16.3% (68,926) | 16.7% (57,706)*** | 11.9% (16,076)*** |
| Morbid Obesity | 8.7% (1,264,568)*** | 9.8% (41,133) | 13.9% (48,083)*** | 9.6% (13,013)*** |

† Asterisks shown in this column indicate p values for the differences between individuals who have not been diagnosed with Covid-19 and confirmed Covid-19 cases in the Salus Medicare cohort.
‡ Covid-19 outpatients are defined as individuals who did not require hospitalization for the disease and remained alive at least 30 days after diagnosis. Covid-19 cases who either did not die or were not hospitalized for the disease but who had no claims more than 30 days after their Covid-19 diagnosis are not shown in this table.
¶ Covid-19 hospitalized cases are those requiring inpatient admission for management of their disease. Asterisks shown in this column indicate the p values for differences between this group and the Covid-19 outpatient group.

Among Covid-19 cases, patients with the most severe disease resulting in either hospitalization or death had higher frequencies of diabetes, COPD, ESRD, chronic kidney disease, hypertension, ischemic heart disease, cerebrovascular disease, pulmonary fibrosis or pulmonary hypertension, chronic liver disease, asthma and congestive heart failure (by Chi Squared tests, all P < 0.001; see Table 1). Patients hospitalized for Covid-19 had similar comorbidity frequencies to those reported by CMS in its monthly Medicare Covid-19 Data Snapshot^14^. With regard to social vulnerability, beneficiaries who were either hospitalized or died due to Covid-19 had higher frequencies of living in zip codes with the lowest income, or highest multiunit housing (by Chi Squared tests, all P < 0.001), or with the most crowded housing (Chi squared tests, all P < 0.01; see Table 1) than patients with less severe disease or beneficiaries who did not have Covid-19.

### Individual Predictors

Logistic regression adjusted odds ratios identifying predictors of hospitalization and death at the individual level are presented in Figures 1 and 2, respectively. The hospitalization model achieved an Area Under the Receiver Operating Characteristic curve (AUROC) of 0.66 (balanced accuracy 0.61 using threshold 0.50) while the death model achieved an AUROC of 0.71 (balanced accuracy 0.65 using threshold 0.50). Variables excluded from the models based on the specified selection criteria to remove insignificant variables are listed in the legends of Figures 1 and 2.

**Figure 1:**
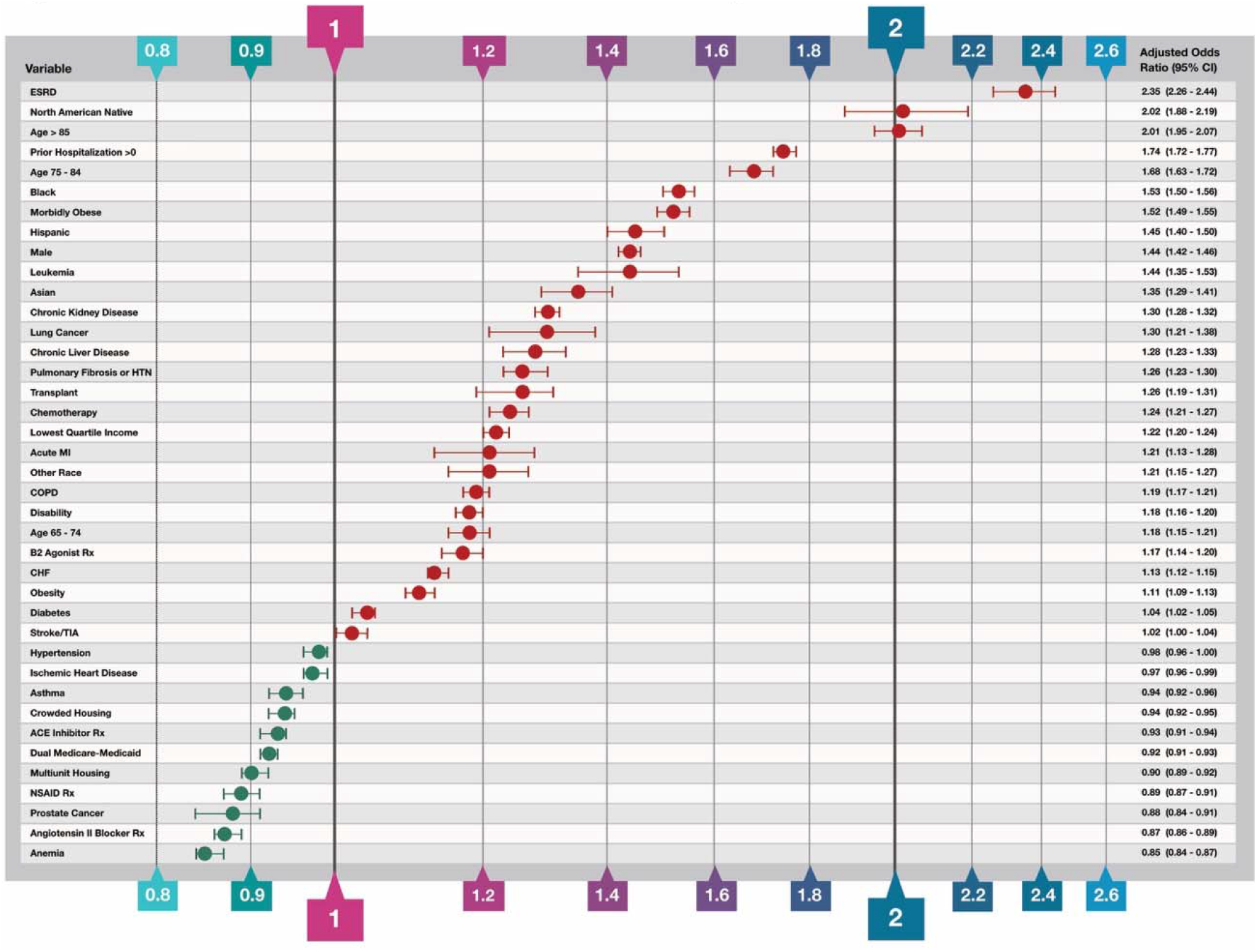
Predictor Variables for Covid-19 Related Hospitalization. The independent variable Odds Ratios were determined by binary logistic regression analysis of confirmed Covid-19 cases that required hospitalization for the disease and for those that were managed with outpatient care only. In addition to the thirty-nine variables shown in the figure, the following variables were included in the model based on the variable selection criteria described in Methods but are not shown: colorectal cancer (OR 1.07; 95% CI 1.01 - 1.14), endometrial cancer (OR 1.12; 95% CI 1.00 - 1.25) in the second half of 2019, other ethnicity (OR 1.19; 95% CI 1.13 - 1.25), unknown ethnicity (OR 0.96; 95% CI 0.91 - 1.00), prescriptions overlapping the Covid-19 diagnosis date of Azithromycin (OR 1.15; 95% CI 1.11 - 1.18), Chloroquine and Hydroxychloroquine drugs (OR 0.96; 95% CI 0.91 - 1.01), anticoagulant drugs (OR 1.06; 95% CI 1.04 - 1.08), opioid drugs (OR 1.03; 95% CI 1.01 - 1.05) and H2 blocker drugs (OR 1.03; 95% CI 0.99 - 1.06); Variables excluded from the model based on the variable selection criteria included: a history breast cancer in the second half of 2019, prescriptions for immunosuppressive and corticosteroid drugs overlapping the Covid-19 diagnosis date, hypertension and pneumococcal vaccinations.

**Figure 2:**
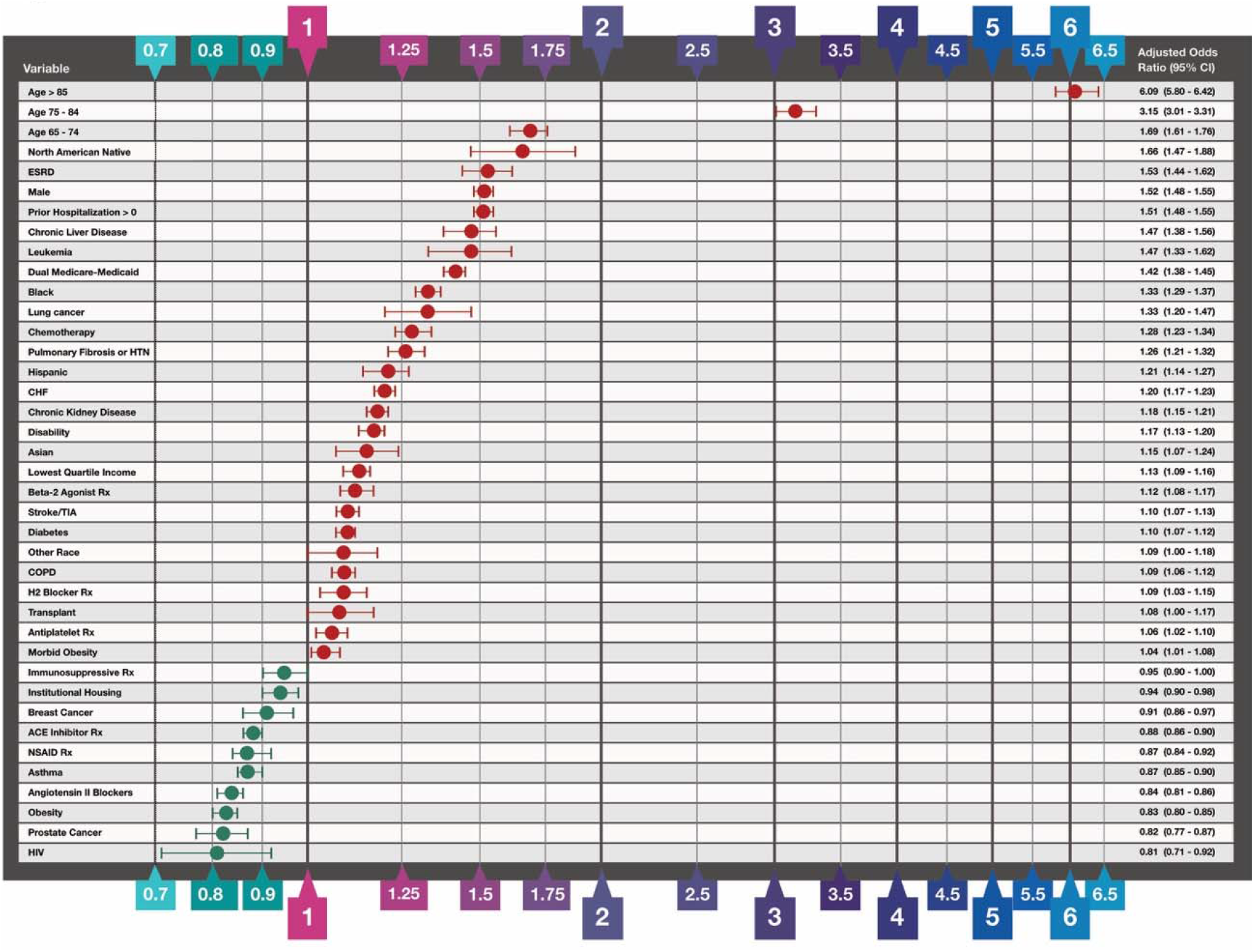
Predictor Variables for Covid-19 Related Death. The independent variable Odds Ratios were determined by binary logistic regression analysis of confirmed Covid-19 cases that survived and those that died within 60 days of Covid-19 diagnosis. In addition to the thirty-nine variables shown in the figure, the following variables were included in the model based on the variable selection criteria described in Methods but are not shown: prescriptions filled with sufficient quantity to overlap the Covid-19 diagnosis date: Azithromycin (OR 1.18; 95% CI 1.13 - 1.23), chloroquine and hydroxychloroquine drugs (OR 1.22; 95% CI 1.13 - 1.23), unknown race (OR 0.88; 95% CI 0.80 - 0.96). Odds ratios for anemia and prescriptions for anticoagulant drugs and corticosteroids had regression coefficient p values > 0.05 and are not shown. Variables excluded from the model based on the variable selection criteria include a history of colorectal cancer and endometrial cancer, or acute MI between July and December 2019, ischemic heart disease, hypertension, residence in zip codes in the top quartile of crowded housing or multiunit housing, and prescriptions for opioid drugs.

As expected, demographic factors are among the strongest predictors of hospitalization and death. The 85+ age group was twice as likely to be hospitalized relative to the reference level (OR 2.20; 95% CI 1.88 - 2.19) and six times more likely to die following a Covid-19 diagnosis (death OR 6.09; 95% CI 5.90 - 6.42). High risk was also observed in other two older age groups: 65-74 (death OR 1.69; 95% CI 1.61 - 1.76) and 75-85 (hospitalization OR 1.68; 95% CI 1.63 - 1.72; death OR 3.15; 95% CI 3.01-3.31). North American Natives were twice as likely to be hospitalized (OR 2.02; 95% CI 1.88 - 2.19) and 66% more likely to die after Covid-19 diagnosis. In addition, Black (hospitalization OR 1.53; 95% CI 1.50 - 1.56) and Hispanic (OR 1.45; 95% CI 1.40 - 1.50) ethnicities were associated with higher risk of hospitalization.

Among socioeconomic factors extracted from SVI data, the strongest predictor for severe Covid-19 was living in a zip code with the lowest quartile of income (hospitalization OR 1.22; 95% CI 1.20 - 1.24; death OR 1.13; 95% CI 1.09 - 1.16). In contrast to income, neither crowded housing nor living in multiunit housing were independent risk factors for either hospitalization or death from Covid-19 (both ORs < 1.0). The socio-economic variables for poverty and education, the housing and transportation summary SVI ranking variable, and the overall summary SVI variable were highly correlated with either income or housing variables and thus excluded from the final models (see online Supplement for more details).

On the clinical side, the strongest risk factors for Covid-19 hospitalization and death are End-Stage Renal Disease (ESRD, hospitalization OR 2.35; 95% CI 2.26 - 2.44, death OR 1.53; 95% CI 1.44 - 1.62) and frailty as assessed by one or more prior hospitalization(s) since October 2019 (hospitalization OR 1.74; 95% CI 1.72 - 1.77; death OR 1.51; 95% CI 1.48 - 1.55). Morbid obesity and leukemia were leading risk factors for hospitalization; in addition, patients with chronic kidney disease, lung cancer, chronic liver disease, pulmonary fibrosis or pulmonary hypertension, or were undergoing chemotherapy had 20-30% higher risk of hospitalization (see Figure 1 for details). Beneficiaries with leukemia (OR 1.47; 95% CI 1.33-1.62) and chronic liver disease (OR 1.47; 95% CI 1.38-1.56) were associated with a 47% higher risk of death following Covid-19 diagnosis. COPD alone was associated with a modest increased risk of hospitalization (OR 1.19; 95% CI 1.17 – 1.21). However, beneficiaries with COPD who also had a drug claim for a beta-2 agonist bronchodilator would have an OR of hospitalization of 1.39 (calculated by adding their respective regression coefficients together) which would put them in the highest risk quartile. On the other hand, diabetes was not a major predictor of hospitalization (OR 1.04; 95% CI 1.02 - 1.05) but diabetic Covid-19 patients were at a slightly higher risk of death (OR 1.10; 95% CI 1.07 - 1.12). Hypertension and asthma were not associated with higher odds of Covid-19 hospitalizations or death.

Use of ACE inhibitors, angiotensin II blockers, and NSAIDs were modestly associated with lower rates of Covid-19-related hospitalization or death.

The Random Forest results identifying important factors for predicting hospitalization and death in the Medicare beneficiary sample are presented in Figure 3. The Random Forest hospitalization model achieved an AUROC of 0.67 while the death model achieved an AUROC of 0.71. The Feature Importance (FI) values for the random forest models are shown in Figure 3, and mainly confirm the regression analysis findings. A history of prior hospitalizations before the diagnosis of Covid-19 was the most important variable in the hospitalization model (FI 0.17), but chronic kidney disease (FI 0.08) and ESRD (FI 0.05) were also identified as important features. The most important predictor of death was age > 85 (FI 0.16), but a history of prior hospitalizations (FI 0.06) and congestive heart failure (0.06) were also important.

**Figure 3:**
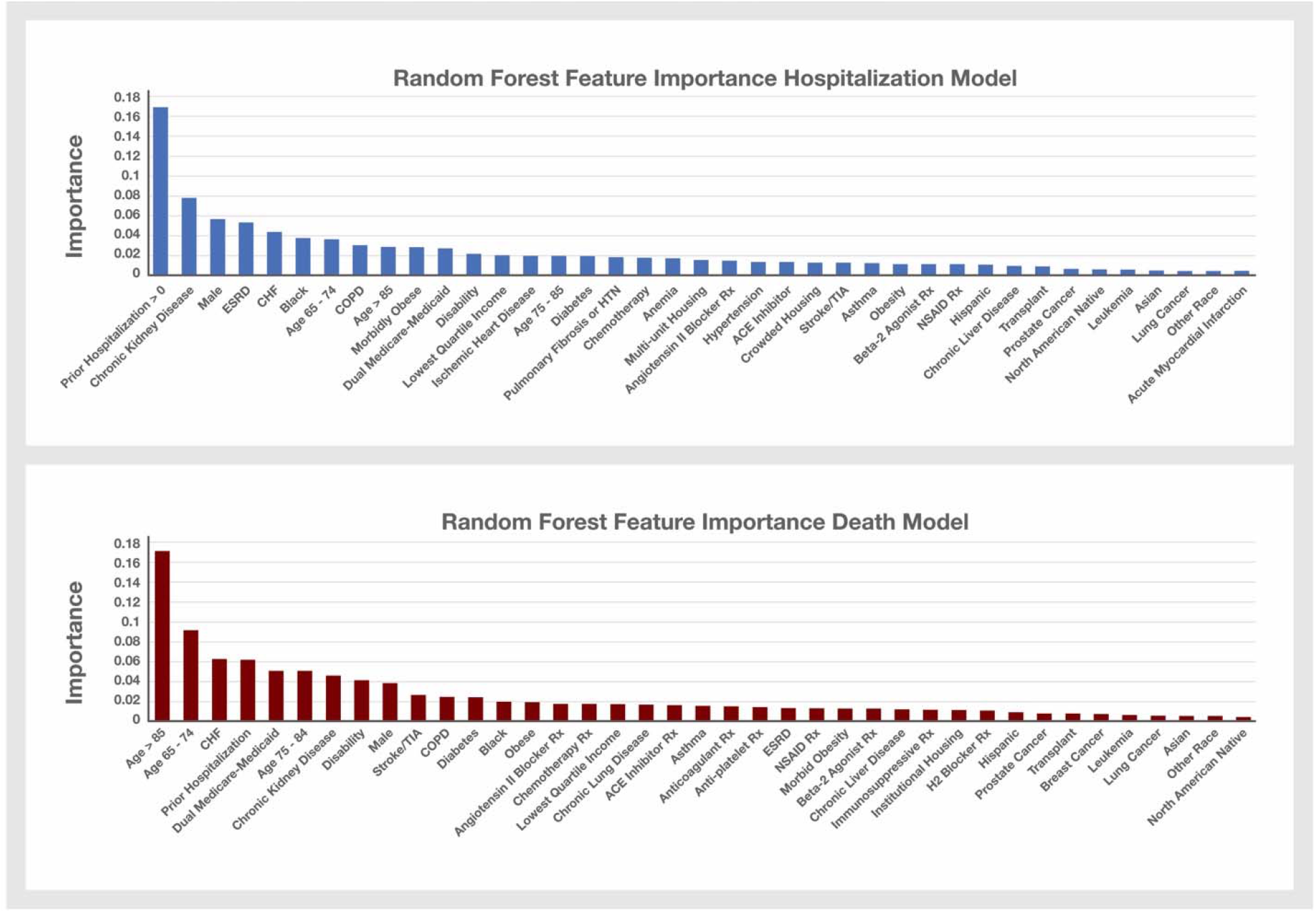
Random Forest Hospitalization and Death Model Feature Importance. Variables that were selected for inclusion in the Hospitalization and Death logistic regression models were used to build these two random forest models. The Feature Importance values for the variables not shown in the Hospitalization model graph are: prescriptions filled with sufficient quantity to overlap the Covid-19 diagnosis date for Azithromycin (FI 0.0104), Chloroquine and Hydroxychloroquine drugs (FI 0.0056), anticoagulant drugs (FI 0.0129), antiplatelet drugs (FI 0.0105), corticosteroids (FI 0.0118), and immunosuppressive drugs (FI 0.100); endometrial cancer (FI 0.002) or breast cancer (FI 0.006) between July and December 2019; unknown race (0.0039) and HIV (0.0045). The Feature Importance values for the variables not shown in the Death model graph include: prescriptions filled with sufficient quantity to overlap the Covid-19 diagnosis date for Azithromycin (0.0107), Chloroquine and Hydroxychloroquine drugs (0.0065), corticosteroids (FI 0.0134), anemia (FI 0.0189), unknown race (FI 0.004) and HIV (FI 0.0037).

### Population Risk Mapping and Stratification

We used the logistic regression results to calculate a predicted probability of hospitalization for every individual in the cohort and applied these probabilities to map population level risk. The risk map displays, with use of a color gradient, the percentage of our cohort with a predicted probability of hospitalization greater than 0.55 for every residential zip code in the U.S.

Figure 4 displays an example of the risk map in the Los Angeles metropolitan area. In this example, the Los Angeles zip codes displaying a wide range of population percentages over the 0.55 predicted hospitalization probability threshold, were positively correlated with the cumulative Covid-19 case hospitalization rates in these zip codes (Pearson’s R = 0.59, (df = 306; p < 0.0001)). We further conducted the same correlation and linear regression analyses in 5 other major metropolitan areas in the U.S. All correlation coefficients in these regions had p values < 0.0001 by Chi Square. (see Figure S1 in the Supplemental Appendix). For the 15 most populous U.S. metropolitan areas, we also observed a positive correlation of risk levels with Covid-19 case hospitalization rates (mean Pearson’s R = 0.51; 95% CI 0.45 – 0.56 with P values ranging between < 0.0001 and 0.03).

**Figure 4:**
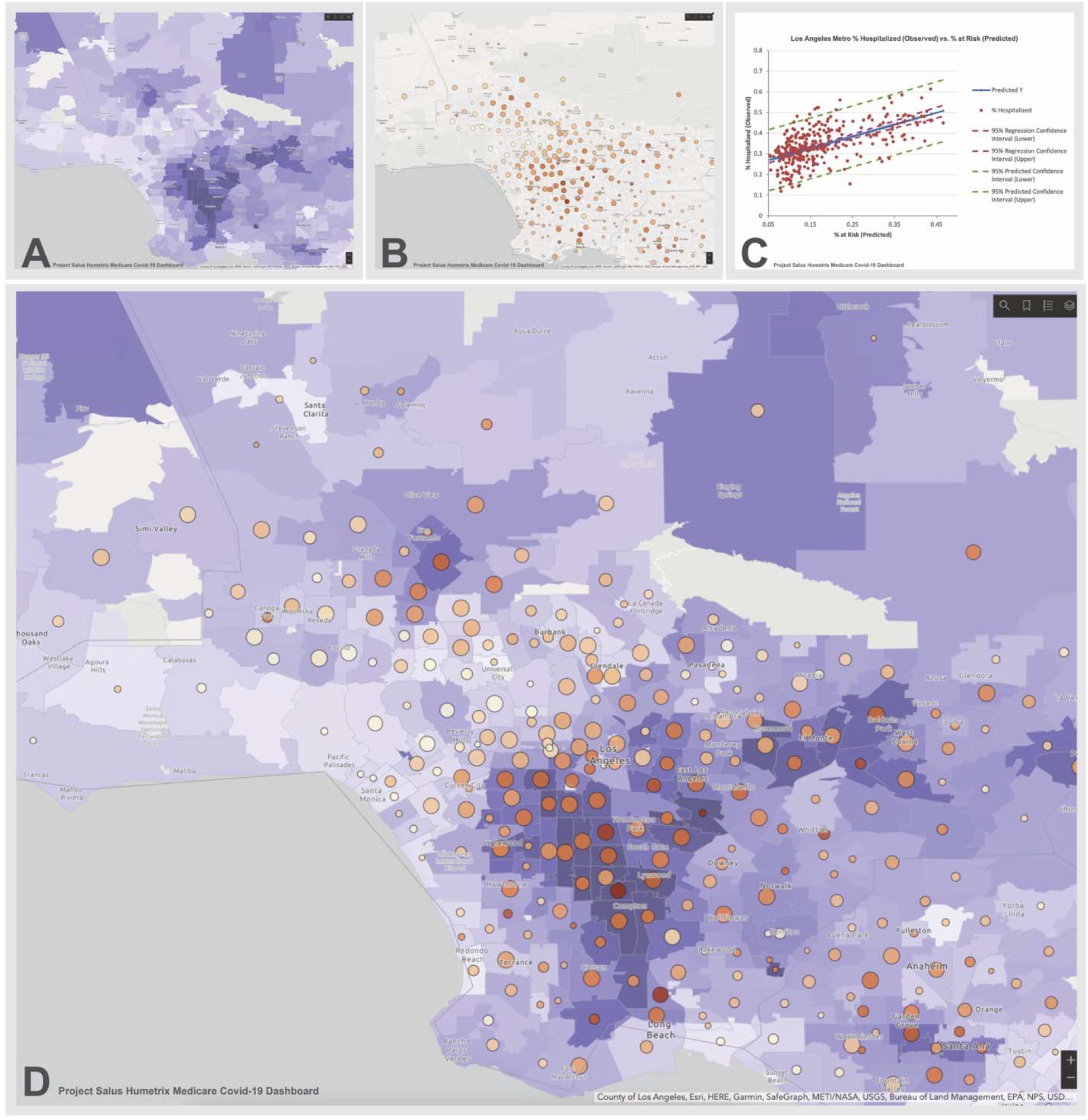
Los Angeles Covid-19 Hospitalization Risk Map. Panel A shows the percentage of the Salus cohort with a predicted probability of hospitalization when diagnosed with Covid-19 of over 0.55 on a light blue to dark lavender color scale. Panel B shows the cumulative number of hospitalizations per zip code (increasing size of circles denotes a higher hospitalization count) with the percentage of cases requiring hospitalization shown on a beige to dark orange scale. Panel C shows a linear regression analysis of the case hospitalization rate (Y axis) as a function of the risk level in each zip code (regression R^2^ = 0.35); Panel D is an overlay of panel B on Panel A and demonstrates that zip codes with the highest predicted probabilities of hospitalization with Covid-19 tend to have higher observed percentage of cases requiring hospitalization and vice versa.

A nationwide risk map generated by the risk model is currently available through the Department of Defense Project Salus.

## DISCUSSION

The severe Covid-19 risk models and the mapping of Medicare beneficiaries at highest risk are based, to our knowledge, on the largest Covid-19 dataset assembled to date for this purpose. Importantly, this cohort captured all confirmed Covid-19 cases, their hospitalizations and deaths amongst the total 37 million beneficiaries comprising the Medicare FFS population. All members of the study cohort were active users of healthcare services, presenting on average a frailer clinical profile than the general Medicare population,^22^ as shown by their clinical characteristics in Table 1. While our observed 1,030,893 Medicare FFS cases represent a little less than 10% of the total number of Covid-19 cases in the U.S., they contribute close to 50% of all CDC estimated Covid-19-related hospitalizations and a majority of Medicare hospitalizations.^23^ The scale of this dataset has allowed us to quantitively and qualitatively validate our hospitalization model predictions with actual Covid-19 case hospitalization rates in multiple geographic areas.

The nationwide distribution of the population also enabled DoD’s Project Salus to produce the first county and zip code level mapping of the population at higher risk for severe Covid-19 and related hospitalizations, with the original goal of providing logistics support to hospitals predicted to experience a surge in patients. Beyond its use for pandemic mapping, the model and related mapping provide information that local health authorities can use to operationalizeCovid-19 vaccine distribution and allocation.

Our combined use of logistic regression and random forest analyses, as applied to this very large Medicare population, allowed us to test the current CDC listing of clinical and socio-demographic risk factors for severe Covid-19. Our different analytic approaches provide two different perspectives: the Odds Ratios from the logistic regression help identify individual risk while the impurity-based Feature Importance from the random forest analyses identified the most important variables for predicting severe Covid-19 outcomes for the entire cohort. For example, in the hospitalization model, the second highest Odds Ratio was associated with North American Native ethnicity. Unlike Odds Ratios which are not influenced by the prevalence of the feature in the population, Feature Importance is more influenced by how common a feature is within the sample. Thus, while North American Native ethnicity has one of the highest Odds Ratios in the hospitalization logistic regression model, its Feature Importance in the random forest model was low (ranked 42nd out of 50 variables) due to the low frequency of this feature in the sample (< 1% of the sample).

Both logistic regression and random forest analyses affirm the critical risk factors of ethnicity^24^, older age, and morbid obesity,^25^ as previously reported. However, contrary to prior descriptive analyses performed on smaller population sizes of hospitalized patients only, our analyses reveal the lack of or modest effect of hypertension, diabetes,^26^ COPD,^27^ and asthma,^28^ in our mostly older Medicare beneficiary population. Both analytical approaches also show that prior hospitalizations, a marker of frailty in aged Medicare beneficiaries,^29^ was one of the most significant individual characteristics associated with severe Covid-19 outcomes.

One of the main limitations of our study and derived models is that it is only based on the Medicare FFS population, which represents approximately 60% of the total Medicare population (with regional variations ranging from 98% in Alaska to 51% in Minnesota). There is evidence that Medicare Advantage plans tend to enroll beneficiaries who are healthier than Medicare FFS beneficiaries, this difference in health status will limit the generalization of our model to the entire Medicare population. If the model were to be used for vaccine allocation, it could be updated using Medicare Advantage data if it were made available.

With the above limitations, the models we have developed provide important information for clinicians and policy makers to consider. Specifically, because the models integrating both socio-economic factors and individual clinical data respond to the recommendations of the NAM for prioritization and allocation of Covid-19 vaccines, they could be used to support planning a vaccination campaign. Figure 5 displays a histogram the distribution of the predicted probabilities of hospitalization for SARS-CoV-2 infected patients, and when such data are mapped, they enable planners to estimate how many high-risk beneficiaries reside in a jurisdiction, and of those, how many are in socially vulnerable areas. Our models identifying individuals at risk for severe Covid-19 could also be used by the Medicare program, in collaboration with state and local health officials, to affirmatively invite or encourage highest risk beneficiaries to seek early vaccination. Additionally, as is done for electricity dependent persons in other emergencies, local health officials can request names and addresses of these beneficiaries in their jurisdictions, provided they can HIPAA-protect the data, and then could conduct direct outreach to beneficiaries at highest risk. Further, once receipt of vaccination is linked to Medicare claims, the analytic platform could be used to support post-licensure pharmacovigilance and effectiveness studies, especially for those identified at higher risk of severe Covid-19. These are of paramount importance, especially in the early phases of a vaccination campaign.

**Figure 5:**
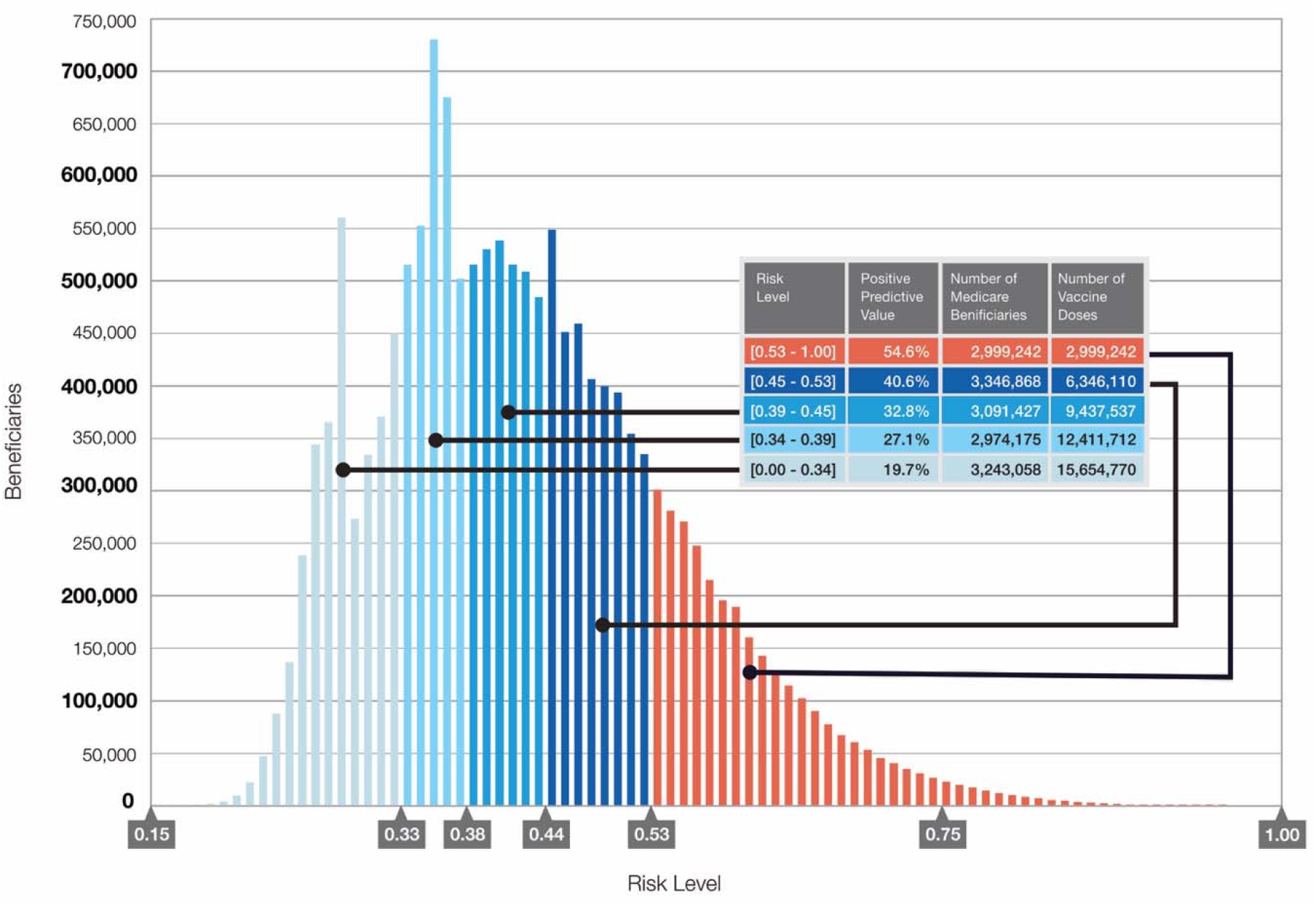
Covid-19 Vaccine Prioritization Based on Risk of Severe Covid-19 Disease. The logistic regression model coefficients for the independent variables shown in figure 1 were used to calculate the predicted probabilities of hospitalization in the Salus cohort. The distribution of predicted probabilities was split into 5 groups shown in the table of approximately 3M beneficiaries each to enable stratification of the cohort by risk of severe disease in order to prioritize individuals for Covid-19 vaccination.

We believe that this quantitative analysis of risk for severe Covid-19 in the Medicare population provides important new insights useful for managing the remainder of the Covid-19 pandemic, including a vaccination campaign. To be immediately actionable, state and local governments could consider asking the DoD, through the usual Mission Assignment process used to provide domestic support, to provide the risk mapping to their jurisdictions. Finally, this work underscores the value of Medicare claim data for epidemiologic surveillance, with its size and nationwide representation, can also augment both the ILINet and COVID-NET disease surveillance systems and complement the existing vaccine monitoring systems for tracking both the safety and efficacy of Covid-19 vaccination in the high-risk Medicare population.

## Data Availability

The Medicare claim data files used in this research is protected health information as defined by the Health Insurance Portability and Accountability Act (HIPAA). Access to these data sets require CMS approval.
The 2018 CDC Social Vulnerability Index data can be downloaded from the CDC website.

https://www.atsdr.cdc.gov/placeandhealth/svi/data_documentation_download.html

## ACKNOWLEDGEMENTS

We thank Robert Lyden, Travis Yatsko, Adrien Cirou, and Franz Krachtus of Humetrix for their assistance in project management and in the production of the manuscript figures and tables.

## CONFLICT OF INTEREST DISCLOSURES

Bettina Experton, Adrien Elena, Chris Hein, Blake Schwendiman and Chris Burrow of Humetrix disclose the following conflict of interest: the study conducted by Humetrix for Project Salus has been funded by the Johns Hopkins University Applied Physics Laboratory under a prime contract with the Department of Defense Joint Artificial Intelligence Center.

Justin Vincent of Amazon Web Services, discloses the following conflict of interest: received fees for professional services rendered for Project Salus from ECS Federal LLC, part of ASGN Incorporated, under a prime contract of the Department of Defense Joint Artificial Intelligence Center.

Nicole Lurie, Hassan Tetteh, Peter Walker and Colin Carroll have no conflict of interest to disclose.

### Department of Defense Disclaimer

The views expressed in this article are those of the authors and do not necessarily reflect the official policy or position of the Department of Defense, nor the U.S. Government. Authors are military service members (or an employee of the US Government). This work was prepared as part of their official duties. Title 17 USC§105 provides that copyright protection under this title is not available for any work of the US Government. Title 17 USC§101 defines a US Government work as a work prepared by a military service member or employee of the US Government as part of that person’s official duties.

## REFERENCES

1. Sara Oliver M.D. Centers for Disease Control and Prevention. ACIP COVID-19 Vaccines Work Group. September 22, 2020. https://www.cdc.gov/vaccines/acip/meetings/downloads/slides-2020-09/COVID-06-Oliver.pdf. Accessed December 2, 2020.

2. Centers for Disease Control and Prevention. Older Adults. Updated Nov. 27, 2020. https://www.cdc.gov/coronavirus/2019-ncov/need-extra-precautions/older-adults.html. Accessed December 2, 2020.

3. Centers for Disease Control and Prevention. People at Increased Risk. Updated Nov. 30, 2020. https://www.cdc.gov/coronavirus/2019-ncov/need-extra-precautions/index.html?CDC_AA_refVal=https%3A%2F%2Fwww.cdc.gov%2Fcoronavirus%2F2019-ncov%2Fneed-extra-precautions%2Fpeople-at-increased-risk.html. Accessed December 2, 2020.

4. The National Academies of Sciences, Engineering, and Medicine. National Academies Release Framework for Equitable Allocation of a COVID-19 Vaccine for Adoption by HHS, State, Tribal, Local, and Territorial Authorities. October 2, 2020. https://www.nationalacademies.org/news/2020/10/national-academies-release-framework-for-equitable-allocation-of-a-covid-19-vaccine-for-adoption-by-hhs-state-tribal-local-and-territorial-authorities. Accessed December 2, 2020.

5. Centers for Disease Control and Prevention. CDC SVI 2018 Documentation. January 31, 2020. https://svi.cdc.gov/Documents/Data/2018_SVI_Data/SVI2018Documentation.pdf. Accessed December 2, 2020.

6. Surgo Foundation. The COVID-19 Community Vulnerability Index (CCVI). https://precisionforcovid.org/ccvi. Accessed December 2, 2020.

7. Kathleen Dooling, M.D.M.P.H. Centers for Disease Control and Prevention. ACIP COVID-19 Vaccines Work Group. September 22, 2020. https://www.cdc.gov/vaccines/acip/meetings/downloads/slides-2020-09/COVID-07-Dooling.pdf. Accessed December 2, 2020.

8. Centers for Disease Control and Prevention. Percent of U.S. Adults 55 and Over with Chronic Conditions https://www.cdc.gov/nchs/health_policy/adult_chronic_conditions.htm. Accessed December 2, 2020.

9. Kathleen Dooling, M.D.M.P.H. Centers for Disease Control and Prevention. Phased Allocation of COVID-19 Vaccines. November 23, 2020.https://www.cdc.gov/vaccines/acip/meetings/downloads/slides-2020-11/COVID-04-Dooling.pdf. Accessed December 2, 2020.

10. Centers for Disease Control and Prevention. People with Certain Medical Conditions. Updated Dec. 1, 2020. https://www.cdc.gov/coronavirus/2019-ncov/need-extra-precautions/people-with-medical-conditions.html. Accessed December 2, 2020.

11. CDC COVID-19 Response Team. Preliminary Estimates of the Prevalence of Selected Underlying Health Conditions Among Patients with Coronavirus Disease 2019 - United States, February 12 - March 28, 2020. Centers for Disease Control and Prevention. 2020;69(13):382–386.

12. Lithander F, Neumann S, Tenison E, et al. COVID-19 in older people: a rapid clinical review. Oxford Academic. 2020;49(4):501–515.

13. Garg S, Kim L, Whitaker M, et al. Hospitalization Rates and Characteristics of Patients Hospitalized with Laboratory-Confirmed Coronavirus Disease 2019 - COVID-NET, 14 States, March 1 -30, 2020. Centers for Disease Control and Prevention. 2020;69(15):458–464.

14. Centers for Medicare & Medicaid Services. Preliminary Medicare COVID-19 Data Snapshot. https://www.cms.gov/research-statistics-data-systems/preliminary-medicare-covid-19-data-snapshot. Accessed December 2, 2020.

15. AI in Defense DoD’s Artificial Intelligence Blog. The JAIC Forges Ahead. May 20, 2020. https://www.ai.mil/blog_05_20_20-the_jaic_forges_ahead.html. Accessed December 2, 2020.

16. HUD USPS Zip Code Crosswalk Files. Office of Policy Development and Research (PD&R). 2020. https://www.huduser.gov/portal/datasets/usps_crosswalk.html. Accessed December 2, 2020.

17. R Core Team (2019). R: A language and environment for statistical computing and graphics. R Foundation for Statistical Computing, Vienna, Austria. https://www.R-project.org/. Accessed December 2, 2020.

18. Frank E Harrell Jr. rms: Regression Modeling Strategies. July 18, 2020. https://CRAN.R-project.org/package=rms. Accessed December 2, 2020.

19. Robin X, Turck N, Hainard A, et al. pROC: an open-source package for R and S+ to analyze and compare ROC curves [Published online March 17, 2011]. BMC Bioinformatics. doi:10.1186/1471-2105-12-77.

20. Friedman J, Hastie T, Tibshirani R. Regularization Paths for Generalized Linear Models via Coordinate Descent. J Stat Softw. 2010;33(1):1–22.

21. Pedregosa F, Varoquaux G, Gramfort A, et al. Scikit-learn: Machine Learning in Python. J Mach Learn Res. 2011;12:2825–2830.

22. Centers for Medicare & Medicaid Services. Prevalence of Chronic Conditions among Fee-for-Service Beneficiaries: 2017. https://www.cms.gov/Research-Statistics-Data-and-Systems/Statistics-Trends-and-Reports/Chronic-Conditions/Chartbook_Charts. Accessed December 2, 2020.

23. Centers for Disease Control and Prevention. COVID-19 Hospitalization and Death by Age. Updated August 18, 2020. https://www.cdc.gov/coronavirus/2019-ncov/covid-data/investigations-discovery/hospitalization-death-by-age.html. Accessed December 2, 2020.

24. Azar K, Shen Z, Romanelli R, et al. Disparities In Outcomes Among COVID-19 Patients In A Large Health Care System In California [Published online May 21, 2020]. Health Aff. doi:10.1377/hlthaff.2020.00598

25. Palaiodimos L, Kokkinidis D, Li W, et al. Severe Obesity, Increasing Age and Male Sex Are Independently Associated with Worse In-Hospital Outcomes, and Higher In-Hospital Mortality, in a Cohort of Patients with COVID-19 in the Bronx, New York. Metabolism. 2020;108(154262).

26. Richardson S, Hirsch J, Narasimhan M, et al. Presenting Characteristics, Comorbidities, and Outcomes among 5700 Patients Hospitalized with COVID-19 in the New York City Area. JAMA. 2020;323(20):2052–2059.

27. Lippi G, Henry B. Chronic Obtrusive Pulmonary Disease Is Associated with Severe Coronavirus Disease (COVID-19). Respir Med. 2020;167(105941).

28. Song J, Zeng M, Wang H, et al. Distinct Effects of Asthma and COPD Comorbidity on Disease Expression and Outcome in Patients with COVID-19 [Published online July 27, 2020]. Allergy. doi:10.1111/all.14517

29. Experton B, Li Z, Branch L, Ozminkowski R, Mellon-Lacey D. The impact of payor/provider type on health care use and expenditures among the frail elderly. Am J Public Health. 1997;87(2):210–216.

