## Supplemental Appendix for "A Predictive Model for Severe Covid-19 in the Medicare Population: A Tool for Prioritizing Scarce Vaccine Supply"

**Supplemental Methods:**

Medicare claim data processing: The Humetrix SaaS platform installed in the SUNet secure classified government network provides automated pre-processing of weekly updates downloaded to the Humetrix SUNet enclave of Medicare Part A inpatient and outpatient, Hospice and SNF, Part B Carrier claims and Part D (PDE) claims data to generate output files containing derived variables used to train logistic regression and random forest models as described below. The platform’s medical terminology service includes a complete collection of AMA CPT-4, FDA NDC, NNPES NPI, ICD-10-CM, CMS Level II HCPCS, and NLM RxNorm codes and provides automated identification grouping of ICD-10-CM codes to identify chronic condition categories and NDC drug code to RxNorm ingredient code mappings to identify pharmaceutical classes of active pharmaceutical ingredients from Medicare claim data.

Logistic Regression Model Training: We used logistic regression to identify significant predictors of Covid-19 related hospitalization or Covid-19 deaths, (R statistical software, version 3.6 with rms, glmnet and pROC packages) using the binary outcomes of those who received outpatient care only (defined as cases that did not require hospitalization or didn’t die at least thirty days after diagnosis) versus either those who were hospitalized for Covid-19, or those beneficiaries whose deaths were attributed to Covid-19 (defined as cases who died of SARS-CoV-2 infection within 60 days of diagnosis).

We divided our sample into training (60%) and validation (40%) sets to develop our final model, randomly allocating cases to training or validation components in a 50:50 ratio. We examined correlation coefficients between independent variables, and used lasso regression, to eliminate correlated or collinear independent variables. We then used stepwise backward variable selection procedure based on the Akaike Information Criterion (AIC) to remove non-significant variables. Computation of the 95% confidence intervals for the coefficient estimates were generated by bootstrapping (2000 repetitions) on the training set.

Random Forest Model Training: We computed Feature Importance (Python, scikit-learn version 0.22.1 with RandomForestClassifier and GridSearchCV packages).  Trees were build using bootstrapping with balanced subsamples. Parameters specifying the maximum depth of the tree and the number of estimators (trees in the forest) were optimized by cross-validated grid-search in the training set.  The data sampling procedure, variable definition, feature engineering, and patient outcome definitions were identical to those described above for Logistic Regression

Model Validation: Model performance for both logistic regression and random forest hospitalization models was measured by the Area Under the Receiver Operating Characteristics (AUROC). The AUROC was computed on a validation set composed of cases that were not used in the training set, with adjustments to give a 60:40 ratio of outpatient to hospitalized cases (which is the observed ratio for hospitalization in the Covid-19 study population). For the logistic regression and random forest death model the AUROC was computed using an 85:15 ratio (which is the observed case fatality ratio in the Covid-19 study population).

Demographic and Insurance Coverage Variables: weekly updates of CMS “Master Beneficiary Summary 2020 File” (MBSF_2020) data were processed, extracting variables: ORIG_REASON_FOR_ENTITLEMENT (Disability), ZIP_CD (residential zip code), YOB, SEX_CODE, RACE_CODE, ESRD_INDICATOR and DUAL_STUS_CD 01-12 (Dual Medicare-Medicaid insurance).

Social Vulnerability Index (SVI) variables associated with zip codes: these were derived from CDC data, which are categorized by census tract. The variables analyzed using binary logistic regression to determine significant predictor variables for hospitalization or death due to Covid-19 included the following SVI variables: the EPL_PCI income variable, the EPL_CROWD (crowded living), EPL_GROUPQ (institutional housing), and EPL_MUNIT (living in multiunit housing); EPL_POV, EPL_NOHSDP (no high school diploma), RPL_THEME1 (summary socioeconomic variable) were all highly correlated with EPL_PCI and excluded on this basis from model training. RPL_THEME4 was excluded based on its correlations with the housing variables EPL_GROUPQ, EPL_CROWD and EPL_MUNIT as well as not surviving AIC based stepwise regression in prior modeling work; RPL_THEMES (the overall tract summary ranking variable) was excluded based on its high correlation with the low-income variable EPL_PCI.

CMS chronic condition variables: These chronic condition codes found in the MBSF_2019 chronic condition segment file were used in hospitalization and death models:

1. Acute myocardial infarction July-December 2019 (AMI)
2. Chronic kidney disease (CHRONIC_KIDNEY_EVER), COPD (COPD_EVER)
3. Congestive heart failure (CHF_EVER)
4. Diabetes mellitus (DIABETES_EVER)
5. Ischemic heart disease (ISCHEMICHEART_EVER)
6. Stroke/transient ischemic attack (STROKE_TIA_EVER)
7. Breast cancer July-December 2019 (CANCER_BREAST)
8. Colorectal cancer July-December 2019 (CANCER_COLORECTAL)
9. Prostate cancer July-December 2019 (CANCER_PROSTATE)
10. Lung cancer July-December 2019 (CANCER_LUNG)
11. Endometrial cancer July-December 2019 (CANCER_ENDOMETRIAL)
12. Anemia July-December 2019 (ANEMIA)
13. Asthma (ASTHMA_EVER)
14. Hypertension (HYPERT_EVER)

Humetrix identified chronic condition variables (present prior to the Covid-19 diagnosis date). The following chronic condition variables were identified by identifying ICD-10-CM codes for the conditions listed below in these CMS claim files: Part A institutional Inpatient and Outpatient claims, Part B Carrier claims, SNF and Hospice claims from October 1, 2019 received by CMS Chronic Condition Warehouse by November 11, 2020.

*Leukemia* (includes acute and chronic myeloid and lymphocytic leukemias as well as less common leukemias); ICD-10-CM codes:

C95, C959, C9590, C9592, C94, C950, C9500, C9502, C92, C929, C9290, C9292, C951, C9510, C9512, C91, C919, C9190, C9192, C93, C939, C9390, C9392, C943, C9430, C9432, C914, C9140, C9142, C901, C9010, C9012, C91A, C91A0, C91A2, C926, C9260, C9262, C92A, C92A0, C92A2, C92Z, C92Z0, C92Z2, C913, C9130, C9132, C916, C9160, C9162, C91Z, C91Z0, C91Z2, C921, C9210, C9212, C93Z, C93Z0, C93Z2, C940, C9400, C9402, C948, C9480, C9482, Z806, C920, C9200, C9202, C910, C9100, C9102, C911, C9110, C9112, C924, C9240, C9242, Z856, C925, C9250, C9252, C915, C9150, C9152, C931, C9310, C9312, C942, C9420, C9422, C933, C9330, C9332, C922, C9220, C9222, C930, C9300, C9302

*Pulmonary fibrosis or pulmonary hypertension* (include idiopathic pulmonary fibrosis, interstitial lung disease, pneumoconioses, pulmonary sarcoidosis, pulmonary hypertension); ICD-10-CM codes:

J61, J62, J620, J628, J63, J636, J65, J64, J60, J84, J841, J8417, J848, J849, J8410, J84112, E84, E840, E841, E8419, E848, E849, I270, I272, I2720, I2722, I2723, I2724, D860.

*Chronic liver disease* (includes alcoholic cirrhosis, primary biliary cirrhosis, chronic viral hepatitis due to hepatitis B and C, alcoholic fatty liver, primary sclerosing cholangitis, Wilson’s disease); ICD-10-CM codes:

K7469, K745, K703, K7030, K7031, P7881, K7460, K74, K743, K744, K746, K717, B18, B180, B181, B182, B188, B189, K70, K709, E8301, K8301, K700, E8801

*HIV/AIDS*; ICD-10-CM codes:

B20, Z717, O9873, O9872, O98719, O98713, O98712, O98711, O9871, O987, B9735, Z21, Z830

*Transplant* (includes following transplants: lung, bone, heart, liver, pancreas, intestine, kidney, bone marrow); ICD-10-CM codes:

Z94, Z949, Z9885, D47Z1, T86810, T86811, T86812, Z942, Z945, Z946, T8621, T8622, T8623, T8641, T8642, T8643, T86890, T86891, T86892, Z941, Z944, Z948, T8611, T8612, T8613, Z940, T86840, T86841, T86842, Z947, Z9483, T86850, T86851, T86852, Z9482, T8631, T8632, T8633, Z9484, T8601, T8602, T8603, T8691, T8692, T8693, Z9481, Z7682, Z943, T86, T8681, T862, T864, T868, T8689, T861, Y830, T8685, T865, T863, T86818, T869, T860, T8619, T8629, T8649, T86898, T86848, T86819, T86858, T8620, T8639, T8640, T86899, C802, T8609, T8610, T8699, T86849, T86859, T8630, Z4824, T8600, T8690, Z482, Z4821, Z4823, Z4822, I2575, I25750, I25751, I25758, I25759, I25811, Z48280, Z4829, Z48290, Z4828, I2576, I25760, I25761, I25768, I25769, I25812, I257

*Obesity* (BMI 30 kg/m^2^ - 40kg/m^2^); ICD-10-CM codes:

Z6830, Z6831, Z6832, Z6833, Z6834, Z6835, Z6836, Z6837, Z6838, Z6839, E6609, E661, E668, E669

*Morbid obesity* (BMI over 40kg/m^2^); ICD-10-CM codes:

Z6841, Z6842, Z6843, Z6844, Z6845, E6601, E662

Medication variables: PDE and Claims Data Files were analyzed by the Humetrix SaaS system to derive the variables listed below. Variables 2-11 were identified by mapping NDC drug product codes to RxNorm ingredient codes belonging to the indicated pharmaceutical classes of drugs. A code of 1 (=True) was only assigned to these variables if a prescription was filled with a sufficient quantity to overlap the date of the first claim with a Covid-19 ICD-10-CM code. For variable 1 (chemotherapy), the same rule applied to PDE claim NDC codes. For variable 12 (Azithromycin and Choloquine/Hydroxychloroquine), we also assigned a code of 1 for a PDE claim date up to 10 days beyond the first claim with a Covid-19 ICD-10-CM code. RxNorm TTY = IN ingredient codes for all pharmaceutical classes available on request.

1. Chemotherapy: signifies that a beneficiary at any time in 2020 either had an ICD10 code for chemotherapy in part A Institutional or Part B Carrier claims, had a HCPCS or CPT-4 code indicating administration of parenteral chemotherapy in a Part B claim, or had a pharmacy (PDE) claim with an NDC code which mapped to an RxNorm ingredient code for an active pharmaceutical ingredient belonging to multiple classes of chemotherapeutic agents based on the National Cancer Institute list of drugs used in the treatment of cancer (<https://www.cancer.gov/about-cancer/treatment/drugs#F>). HCPCS, CPT-4, ICD-10-CM codes are shown below.

CPT-4 chemotherapy codes:

96401, 96402, 96405, 96406, 96409, 96411, 96413, 96415, 96416, 96417, 96420, 96422, 96423, 96425, 96440, 96446, 96450, 96542, 96549, G0498

HCPCS chemotherapy codes:

J8501, J8510, J8515, J8520, J8521, J8530, J8560, J8562, J8565, J8600, J8610, J8650, J8700, J8705, J8999, J9000, J9010, J9015, J9017, J9019, J9020, J9022, J9023, J9025, J9027, J9032, J9033, J9034, J9035, J9036, J9039, J9040, J9041, J9042, J9043, J9044, J9045, J9047, J9050, J9055, J9057, J9060, J9065, J9070, J9098, J9100, J9119, J9120, J9130, J9145, J9150, J9151, J9153, J9160, J9171, J9173, J9176, J9178, J9179, J9181, J9185, J9190, J9199, J9200, J9201, J9203, J9204, J9205, J9206, J9207, J9208, J9210, J9211, J9212, J9213, J9214, J9215, J9216, J9228, J9229, J9230, J9245, J9250, J9260, J9261, J9262, J9263, J9264, J9265, J9266, J9267, J9268, J9269, J9270, J9271, J9280, J9285, J9293, J9295, J9299, J9300, J9301, J9302, J9303, J9305, J9306, J9307, J9308, J9309, J9310, J9311, J9312, J9313, J9315, J9320, J9325, J9328, J9330, J9340, J9351, J9352, J9354, J9355, J9356, J9357, J9360, J9370, J9371, J9390, J9395, J9400, J9600, J9999

ICD-10-CM chemotherapy codes:

D61810, Z5112, Z511, D6481, Z5111, Z511, D701, T80810, T80810D, T80810S, T80810A, D701, D702

2. Anticoagulant drugs (VKORC1 and factor X inhibitors)
3. Antiplatelet drugs (cyclooxygenase inhibitors, ADP receptor inhibitors, adenosine reuptake inhibitors, phosphodiesterase inhibitors)
4. Beta-2 agonists
5. Corticosteroids (excludes topical and ophthalmic preparations)
6. Opioid drugs
7. Histamine type-2 receptor blockers
8. Angiotensin converting enzyme inhibitors (ACE inhibitors)
9. Angiotensin II receptor blockers
10. Non-steroidal anti-inflammatory (NSAID) drugs
11. Immunosuppressive drugs of diverse pharmaceutical classes including anti-interleukins, anti-TNFa drugs, JAK kinase inhibitors, anti-interferons, Sphingosine 1-p receptor modulators, calcineurin inhibitors, mycophenolate and sirolimus and methotrexate.
12. Azithromycin and Chloroquine drugs (includes both Chloroquine and Hydroxychloroquine)

Covid-19 outcome variables:

1. Covid-19 hospitalizations were identified either by Part B Carrier claims with place of service code = 21 or CPT codes indicating inpatient care with a date of service no more than 14 days after or 10 days before the Covid-19 diagnosis date, or by finding Part A Inpatient claims where the data of admission no more than 14 days after or 10 days before the Covid-19 diagnosis date.

CPT-4 codes indicating inpatient services

99217, 99218, 99219, 99220, 99221, 99222, 99223, 99224, 99225, 99226, 99231, 99232, 99233, 99234, 99235, 99236, 99238, 99239

2. Covid-19 cases managed by outpatient care only were defined as cases who were not hospitalized in the 30 days after the Covid-19 diagnosis date and who did not die within 60 days of the diagnosis date.
3. Deaths due to Covid-19 were defined as either a death which occurred during a Covid-19 hospitalization using the inpatient PTNT_DSCHRG_STUS_CD code, or a death which occurred within 60 days of the Covid-19 diagnosis as reported in the DEATH_DT field of the MBSF_2020 file.

**Supplemental Results:**

Effect size calculation (Table 1): Due to the very large sizes of the compared populations, the p values for the statistical tests presented in Table 1 are almost all very significant. We used Cramér’s V to calculate effect sizes values for the comparisons between groups for binary and categorical variables (age ranges, sex, race, income, poverty, dual Medicare-Medicaid dual status, disabled status, prior hospitalization, and clinical variables) all of which had a computed Cramér’s V values under 0.17 indicating a small effect size. We used Vargha and Delanay’s A (VDA) to calculate effect sizes for comparisons between groups for the quantitative variable of age in years and found VDA values of 0.45 (patients with Covid-19 versus those without), 0.55 for those hospitalized for Covid-19 versus those managed as outpatients, and 0.66 for those who died from the disease versus survivors all indicative of small effect sizes.

Analysis of Hospitalization model predictions of Covid-19 case hospitalization rates in six large metropolitan areas (see following pages).

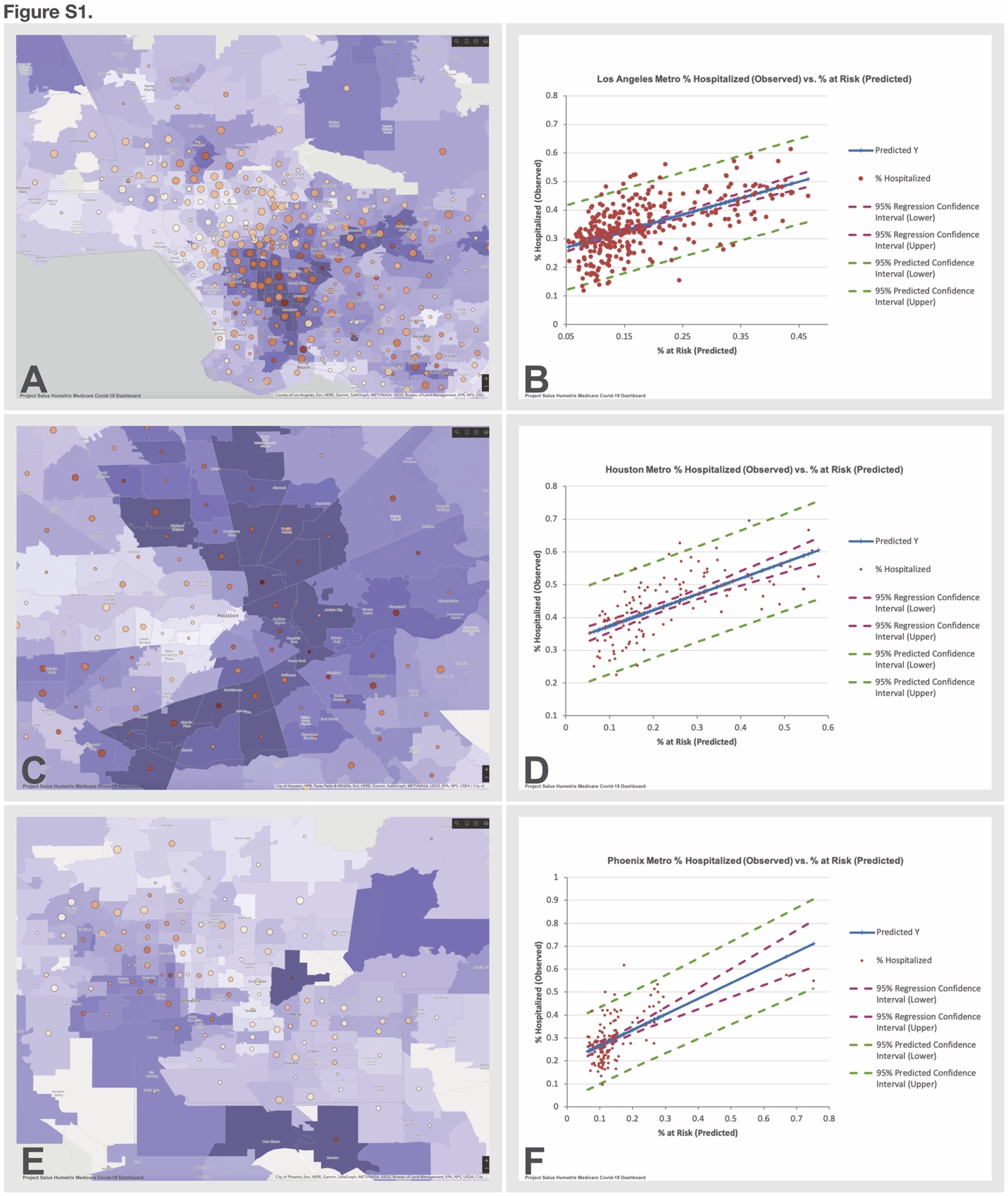

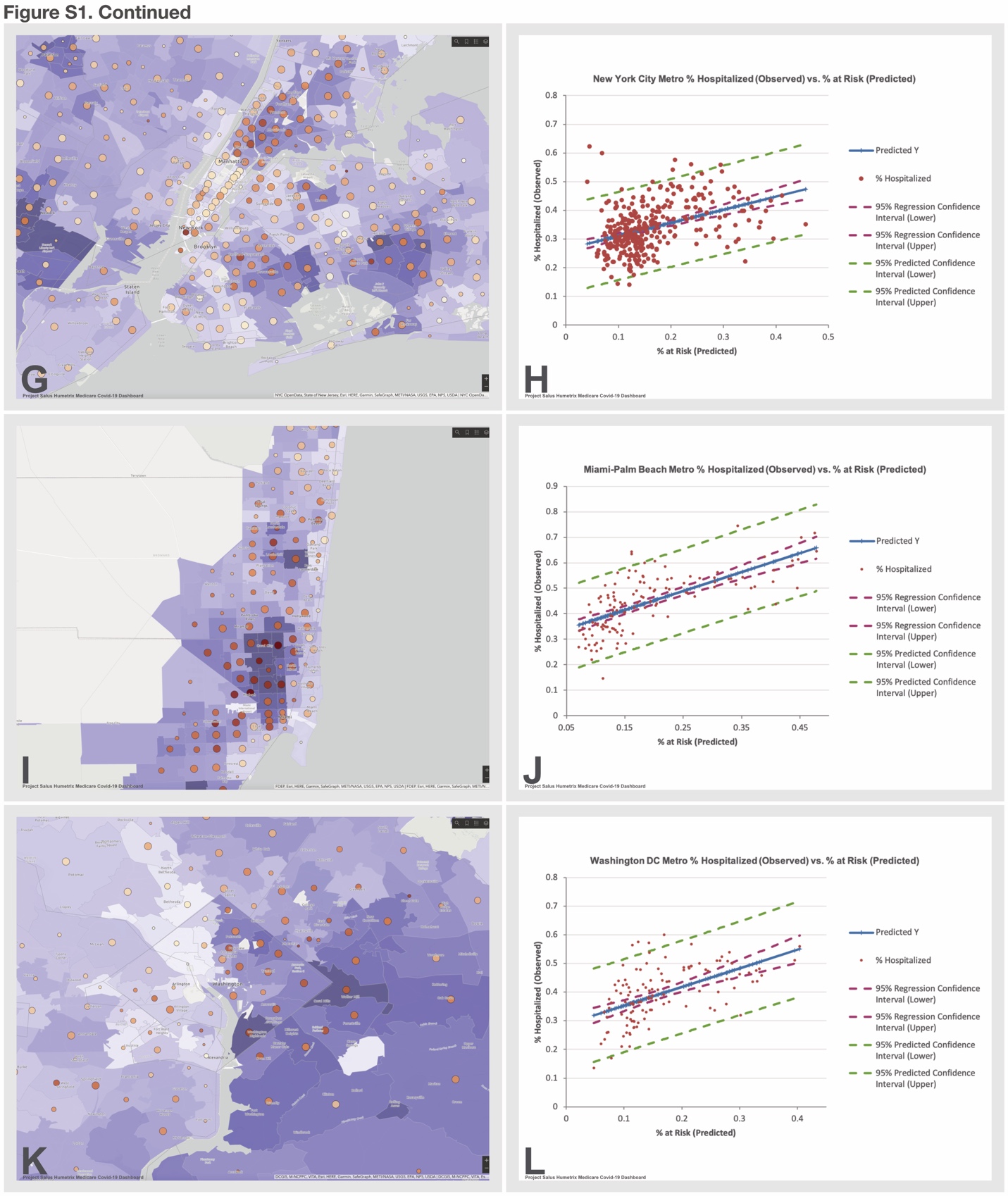

Figure S1: Metropolitan region risk maps for severe Covid-19 (A) Los Angeles County, (C) Houston, (E) Phoenix, (G) New York City, (I) Miami-Dade - Palm Beach, (K) Washington DC. In each metropolitan region, zip codes with the higher predicted probabilities of hospitalization with Covid-19 based on the logistic regression hospitalization model are shown in darker shades of lavender. Circles denote observed cumulative hospitalizations due to Covid-19 extracted from claims data with the size of the

Figure S1 legend continued:

circles scaled to the number of hospitalizations. The percentage of cases requiring hospitalization is displayed by the color of the circles on a beige to dark orange-red color scale with the darker circles indicating higher zip codes with higher percentages of cases requiring hospitalization for Covid-19. Panels B, D, F, H, J, L shows corresponding linear regression analyses of the case hospitalization rates (Y axis) as a function of the risk level in each zip code in each metropolitan region. The following R^2^ values for the regressions were: Los Angeles (0.35), Houston (0.44), Phoenix (0.36), New York City (0.16), Miami-Dade-Palm Beach (0.46), Washington DC (0.29). Metro regions zip codes analyzed available on request.
